# “Immunogenetics of resistance to SARS-CoV-2 infection in discordant couples”

**DOI:** 10.1101/2021.04.21.21255872

**Authors:** Erick C. Castelli, Mateus V. de Castro, Michel S. Naslavsky, Marilia O. Scliar, Nayane S. B. Silva, Heloisa S. Andrade, Andreia S. Souza, Raphaela Neto Pereira, Camila F. B. Castro, Celso T. Mendes-Junior, Diogo Meyer, Kelly Nunes, Larissa R. B. Matos, Monize V. R. Silva, Jaqueline T. W. Wang, Joyce Esposito, Vivian R. Coria, Raul H. Bortolin, Mario H. Hirata, Jhosiene Y. Magawa, Edecio Cunha-Neto, Verônica Coelho, Keity S. Santos, Maria Lucia C. Marin, Jorge Kalil, Miguel Mitne Neto, Rui M. B. Maciel, Maria Rita Passos-Bueno, Mayana Zatz

**Author notes:** Corresponding authors: Mayana Zatz), Erick C. Castelli, Mateus V. de Castro. These two authors contributed equally to this study (Erick C. Castelli, Mateus V. de Castro, Mayana Zatz). **Conflict of interest** The authors declare no conflict of interest.

## Abstract

**Background:** Despite the high number of individuals infected by SARS-CoV-2 who develop COVID-19 symptoms worldwide, many exposed individuals remain asymptomatic and/or stay uninfected. This could be explained by a combination of environmental (exposure, previous infection), epigenetic, and genetic factors. Aiming to identify genetic variants involved in SARS-CoV-2 resistance, we analyzed 86 discordant Brazilian couples where one was infected and symptomatic while the partner remained asymptomatic and seronegative despite sharing the same bedroom during the infection. The discordant partners had comparable ages, and genetic ancestry proportions.

**Methods:** Whole-exome sequencing followed by a state-of-the-art method to call genotypes and haplotypes across the highly polymorphic MHC and LRC.

**Results:** We observed a minor impact in antigen-presentation genes and KIR genes associated with resistance. Interestingly, genes related to immune modulation, mainly involved in NK cell killing activation/inhibition harbor variants potentially contributing to infection resistance. We hypothesize that individuals prone to produce higher amounts of MICA (possibly soluble), LILRB1, LILRB2, and low amounts of MICB, would be more susceptible to infection.

**Conclusion:** According to this hypothesis, quantitative differences in these NK activity-related molecules could contribute to resistance to COVID-19 down regulating NK cell cytotoxic activity in infected individuals but not in resistant partners.

## Introduction

COVID-19, caused by SARS-CoV-2 infection, became a worldwide pandemic affecting millions of people and the leading cause of death in Brazil in 2020 and early 2021. Clinical manifestations range from severe cases with a lethal outcome to mild forms or asymptomatic cases. About 30% of infected individuals are asymptomatic, and they are believed to promote about 60% of transmissions ^1^. Identifying asymptomatic individuals or resistant to the infection who are serum negative is challenging since controlling for and measuring different degrees of exposure is complex. These individuals, however, may provide clues on the mechanisms of resistance and infection itself.

The Human Major Histocompatibility Complex (MHC) is a likely candidate in studying the immune response against infectious agents, such as SARS-CoV-2. It harbors many genes involved in antigen processing (e.g., *TAP1 and TAP2*), presentation (e.g., *HLA-DRB1*), and immune modulation (e.g., *HLA-G, HLA-E, MHC class I–related chain (MIC) A and MICB)*. HLA genes are among the most polymorphic genes in the human genome. *MICA* and *MICB* are also highly polymorphic, with low expression levels in normal tissues but induced in stress situations such as tumors and infections^2^. These proteins differ in binding affinities for its activating receptor NKG2D, present on NK cells, CD8αβ and γδ T cells^3^. Recent studies on the frequency and distribution of HLA alleles and their clinical relevance for the SARS-CoV-2 infection ^4–7^ show inconsistent results, i.e., different associations in different cohorts.

Another natural candidate is the Leukocyte Receptor Complex (LRC). This region encodes killer-cell Ig-like-receptors (KIRs), leukocyte Ig-like receptors (LILRs), and leukocyte-associated Ig-like receptors (LAIRs). The molecules encoded in the LRC have been reported to play important tissue homeostatic functions, affecting innate immune response in stress and the activation and modulation of the adaptive immune response ^8^.

The KIR receptors are expressed on NK cells and some cytotoxic T cells ^9^. KIR genes are highly polymorphic in sequence content, and they present copy number variations. They bind mainly MHC class I molecules and different KIRs recognize different HLA allotypes. These characteristics lead to complex and heterogeneous NK responses, with implications in viral infections, autoimmune diseases, and neoplasias ^10^. LILRs have inhibitory (LILRB) and activatory (LILRA) functions ^8^. They bind mostly to MHC class I molecules and are more conserved than KIR genes. Since viral pathogens can use LILRs as targets to evade the immune responses ^11^, they are also related to responses against viral infections ^12^. The leukocyte-associated Ig-like receptors 1 (LAIR-1) bind collagenous molecules and present inhibitory signals, while LAIR-2 inhibits the interaction between collagen and LAIR-1 ^13^.

Because of the unusually high polymorphism of HLA and KIR loci and their extensive paralogy, these genes are usually neglected in genome-wide association studies (GWAS). Aiming to investigate the variability harbored by genes from the MHC and LRC, and their role in SARS-CoV-2 infection and resistance, we have analyzed a cohort of couples discordant for the infection: one was infected while the household-sharing partner, despite being closely exposed during the infection period, remained asymptomatic and serum negative up to six months afterward. To evaluate the association of variants within the MHC and LRC regions, we whole-exome sequenced these couples and applied a specific bioinformatics pipeline to recover and properly analyse highly polymorphic genes.

## Subjects and Methods

### Volunteers recruitment and datasets

Initial recruitment for screening involved broad media advertising based on the main inclusion criteria: couples discordant for SARS-CoV-2 infection. From more than 1000 received emails, we recruited 100 couples, all from São Paulo (Brazil’s largest metropolis). All couples filled an online questionnaire, which included basic personal information (age, sex, blood type, comorbidities), and clinical progression of COVID-19 as well as diagnostic tests. The non-infected member remained in close contact with his/her symptomatic partner throughout the SARS-CoV-2 infection, sharing the same bed (except when the infected one needed to be hospitalized). Confirmatory tests (RT-PCR for symptomatic and RT-PCR or serology for asymptomatic) endorsed that just one of the pair had symptomatic viral infection at the time. The selected couples were invited to come to the Human Genome and Stem Cell Research Center for an interview and collection of peripheral blood as well as repeated serology assays. The collection of biological samples occurred at intervals from 30 to 180 days after the reported viral infection. Serological testing was repeated in the collected blood plasma with two different techniques (Electrochemiluminescence *and ELISA - SARS-CoV-2 RBD/NP IgA and IgG*). This allowed excluding 7 couples where the asymptomatic partner was found to have IgA or IgG antibodies against SARS-CoV-2. With these criteria, and after sequencing quality control, we report the observed results in 86 selected couples. We also compared the resistant and the infected groups with a previously whole-genome sequenced population-based sample of elderly individuals from the same city ^14^.Importantly, the samples were collected between June and October 2020, before SARS-CoV-2 new variants were reported in Brazil (especially P.1).

### Exome sequencing and variant calling

We used the Nextera Rapid Capture Custom Enrichment Kit or the Nextera Flex Kit (Illumina, San Diego, CA, USA) for library preparation and the IDT xgen-V1 kit for capture following manufacturer protocols. Whole-exome sequencing was performed on the NovaSeq 6000 equipment (Illumina, USA) with a 150 base paired-end dual index read format. Reads were aligned to the human reference GRCh38 using BWA (https://github.com/lh3/bwa/tree/master/bwakit). We also called genotypes using GATK HaplotypeCaller (version 4.0.9). The pipeline used for alignment, variant calling, variant refinement, and genetic ancestry assessment is detailed in the supplementary methods.

### Genotype and haplotype calls for the MHC and LRC

Genes from the MHC and LRC are prone to alignment bias and genotyping errors because of the significant similarity and high polymorphism. To circumvent this issue, we used hla-mapper version 4 ^15^ to optimize read alignment for specific genes from the MHC and LRC (Figure 1), as described elsewhere ^14,15^. After applying hla-mapper, we called genotypes using GATK HaplotypeCaller, with a further refinement step using vcfx. To calculate the copy number of genes in the LRC (Figure 1B), we observed the ratio between the read depths in all exons of each gene and two genes used as references, *TNF* and *LTB*.

**Figure 1:**
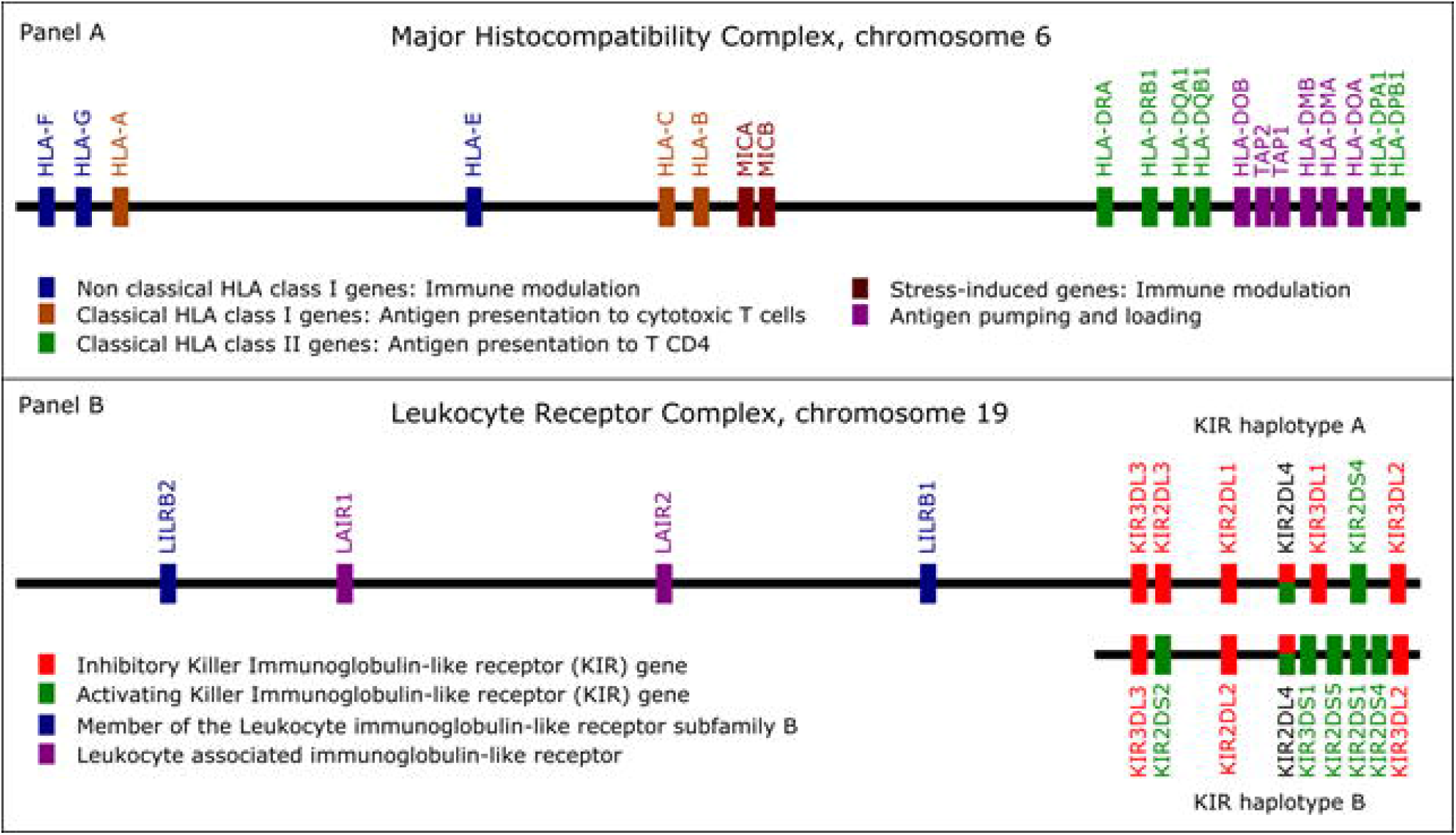
List of the genes optimized by the hla-mapper program. Panel A: The Major Histocompatibility Complex (MHC) and the genes included in the hla-mapper pipeline. Panel B: The Leukocyte Receptor Complex (LRC) and the genes included in the hla-mapper pipeline.

To get phased variants for each gene, we used a strategy described in previous articles addressing HLA diversity in Brazilian samples ^14^, based on both read-aware phasing and probabilistic models. First, we detected the phase between close variants using GATK ReadBackedPhasing. Then, these phase sets were considered by the phasex program.

After the hla-mapper optimization, we obtained accurate genotypes, the complete exonic sequences for each individual, and the translation of these sequences (Figure SM1). These methods are detailed in the supplementary methods, including the procedure to call HLA and KIR alleles.

### Statistical analyses

Association of phenotypic status with bi-allelic and multi-allelic variants was conducted by fitting a logistic regression that considers each allele of a variant as an independent marker, controlling for age, sex, and genetic ancestry, using R and qqman library. Due to sample size limitations, likely multifactorial inheritance, and the high number of variable sites (many multi-allelic) in the MHC and LRC regions, we did not expect large effect sizes to be detected. Therefore, we used a different threshold to detect candidates (P<10-2) that may influence infection susceptibility. To test if rare variants of larger effects contribute to the outcome, we have performed gene-based variant collapsing tests using SKAT-O within the rvtest program ^16^, enabling the analysis of multiallelic variants. SKAT-O is an optimized method for rare variants that combines and compares burden and SKAT tests, resulting in an optimal *P*-value for a given variant set (gene or gene set), controlling for the same covariates as individual variant associations.

We also translated (*in silico*) all exonic sequences into full-length proteins or allotypes (Figure SM1) and evaluated each allotype and amino acid residue by fitting a logistic regression in R, using the same *P*-value threshold and covariates.

## Results

The Resistant and Infected groups had comparable ages, socio-economic status, and genetic ancestry proportions (Table SM1). However, we observed a large sex difference between the two groups. There were 53 men and 33 women among symptomatic individuals, compared to 29 males and 57 females among resistants.

Most of the KIR genes within the LRC presented variation in the number of copies. Thus, we compared the frequency of individuals carrying at least one copy of each gene. The proportion of CNV carriers is similar between the Resistant and Infected groups (Table SR1). Most of the genes presented copy number variation, except *LAIR1, LAIR2, LILRB1, LILRB2, KIR2DL4, KIR3DL2*, and *KIR3DL3*, which have two copies and were considered in SNP-level association analyses.

Here, we evaluated how the MHC and LCR variants may influence susceptibility to SARS-CoV-2 infection. Figure 2 presents the candidate variants for MHC and LCR with *P*-values < 0.01, with only a few variants despite the high number of variants tested (1,723 for the MHC and 632 for the LRC). Because there is high linkage disequilibrium (LD) along the MHC and LRC regions, and some of the associations presented in Figure 2 are not independent, we grouped all variants presenting high LD (r^2^ > 0.9) as haplotypes. Regarding MHC, we found candidate variants for *HLA-C, MICA, MICB, HLA-DRB1*, and *HLA-DOB* genes, including three missense variants in *MICB, HLA-DOB*, and *HLA-DRB1* genes. Among these, only *HLA-DRB1 and HLA-C* molecules have antigen-presenting capacity. Seven out of the 12 candidates are in *MICA* and *MICB*, which are 80 Kb apart. For the LRC region, we detected significant frequency differences for variants at *LILRB1, LILRB2*, and *LAIR2*. The strongest signal coincides with a missense mutation in *LILRB2* (rs7247025-C, *P* = 0.0007), which was overrepresented in infected individuals compared to the general population (*P* = 0.0258), and many variants from *LILRB1* in high LD.

**Figure 2:**
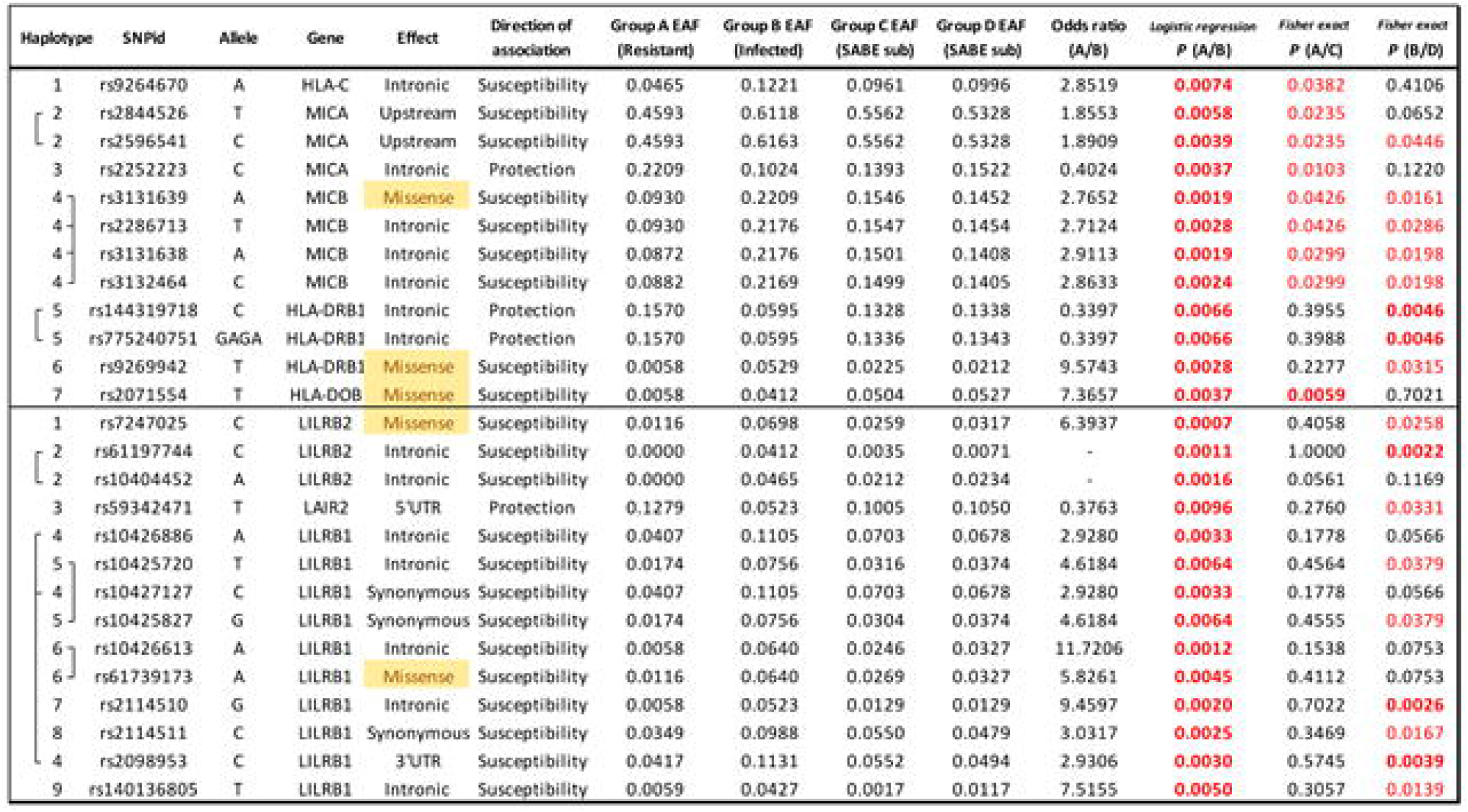
Candidate variants at Major Histocompatibility Complex (MHC) and Leukocyte Receptor Complex (LRC) associated with susceptibility or resistance to SARS-CoV-2 infection (*P* < 0.01). In yellow: missense variants. In bold red, *P*-values below 1%. In red, P-value between 1-5%. Group A: SARS-CoV-2 resistant individuals from the cohort of couples, N = 86. Group B: SARS-CoV-2 infected individuals from the cohort of couples, N = 86. Group C: The frequency in a subsample of the SABE cohort, paired to Group A by genetic ancestry and sex, N = 430. Group D: The frequency in a subsample of the SABE cohort, paired to Group B by genetic ancestry and sex, N = 430. EAF: Effect allele frequency. *P (A/B):* Logistic regression comparing groups A and B, controlled for age, sex, and genetic ancestry. *P (A/C) and P(B/D):* Fisher exact test comparing groups A and C, and B and D.

We also tested how each variant deviates from the expected frequencies when considering a previously collected population sample from the same city and with the same ancestry background and sex (Figure 2). Most of the candidate variants presented intermediate frequencies in the general population samples (groups C and D) when compared to the resistant (Group A) and infected groups (Group B), Figure SR2. This finding may be explained by the mix of resistant and susceptible individuals in the general population.

We applied SKAT-O in each candidate gene to evaluate the contribution of rare and common variants collapsed into gene-wide sets. Among MHC genes, we identified a significant association between *MICB* and infection susceptibility (*P* = 0.011). For the LRC, *LAIR2* was significant (*P* = 0.04). However, we verified that the association signal was driven by the same common variants identified by the single variant association test described in Figure 2.

To investigate how specific protein sequences or amino acid residues may influence susceptibility to SARS-CoV-2 infection, we translated all the exonic sequences to proteins, *in silico*, comparing the frequency of every full-length protein between infected and resistant individuals (Table 1). We also evaluated the frequency distribution of every amino acid residue between groups, but we considered only the genes without copy number polymorphism for this analysis. *MICB* protein sequence named MICB*004:01 and the amino acid residue that defines it, 75-N (related to the missense mutation rs3131639/A), and the protein sequence named HLA-DOB*01:02 and the residue that defines it, 18-Q (related to the missense mutation rs2071554/T), were both associated with susceptibility to infection (Table 1). The associations for *MICB* and *HLA-DOB* remained significant even after correction for multiple tests within each locus. There is also a higher frequency of 101-K at HLA-DRB1 among infected individuals (related to rs9269942/T). We detected one specific protein sequence from *LILRB1* and *LILRB2* (here named Protein-06 and Protein-11, respectively), and the amino acids that define them, 350-R for LILRB1 and 349-G for LILRB2, associated with susceptibility to infection (Table 1). These amino acid exchanges are related to the missense mutations described in Figure 2. Overall, the protein and amino acid analyses corroborate what was observed in the SNP analysis for missense mutations.

**Table 1:** The list of allotypes or amino acid residues from genes from the the Major Histocompatibility Complex (MHC) and the Leukocyte Receptor Complex (LRC) associated with susceptibility or resistance to SARS-CoV-2 infection when controlling for genetic ancestry, sex, and age. Group A: SARS-CoV-2 resistant individuals from the cohort of couples, N = 86. Group B: SARS-CoV-2 infected individuals from the cohort of couples, N = 86.

| Gene | Allotype or amino acid | Association direction | Group A (Resistant) | Group B (Infected) | Odds ratio | P-value |
| --- | --- | --- | --- | --- | --- | --- |
| MICB | MICB*004 | Susceptibility | 0.0814 | 0.2093 | 2.9876 | 0.0014 |
| HLA-DOB | DOB*01:02 | Susceptibility | 0.0058 | 0.0407 | 7.2545 | 0.0037 |
| MICB | 75-N | Susceptibility | 0.0930 | 0.2209 | 2.7650 | 0.0019 |
| HLA-DOB | 18-Q | Susceptibility | 0.0058 | 0.0407 | 7.2581 | 0.0037 |
| HLA-DRB1 | 101-K | Susceptibility | 0.1047 | 0.2151 | 2.3448 | 0.0024 |
| LILRB1 | Protein-6 | Susceptibility | 0.0058 | 0.0465 | 8.3595 | 0.0054 |
| LILRB2 | Protein-11 | Susceptibility | 0.0000 | 0.0417 | 15.6344 | 0.0010 |
| LILRB1 | 350-R | Susceptibility | 0.0116 | 0.0640 | 5.8111 | 0.0045 |
| LILRB2 | 349-G | Susceptibility | 0.0118 | 0.0714 | 6.4619 | 0.0029 |

We also tested *HLA-A* and *HLA-B* alleles grouped as supertypes ^17^. There is a trend to lower frequency of supertype A03 among resistant individuals (*P* = 0.0324) when controlled for genetic ancestry, sex, and age, but this association was not significant after correction for multiple tests (six different HLA-A supertypes).

Finally, we investigated potential mechanisms underlying the associations presented in Figure 2 and Table 1 (detailed at the supplementary results). In brief, the susceptibility-associated variants from *MICA, LILRB1*, and *LILRB2* are associated with higher mRNA expression levels or higher protein stability. In comparison, the susceptibility variants for *MICB* are associated with lower mRNA expression. The susceptibility variant from *HLA-DOB* modifies the signal peptide and possibly the molecule cellular localization and exportation.

## Discussion

Environmental factors such as the use of protective masks and protective measures, socio-economic status, and access to healthcare may explain in part the high variability in COVID-19 disease incidence and mortality among individuals. However, few studies have focused on the genetics of resistance against SARS-CoV-2 infection due to limitations in controlling for exposure. Previous reports on host genetic factors with resistance to COVID-19 have investigated SARS-CoV-2 receptors such as the angiotensin-converting enzyme 2 (ACE2), the transmembrane protease serine-type 2 (TMPRSS2), glucose-regulated protein 78-kDa (GRP78), and the extracellular matrix metalloproteinase inducer or cluster of differentiation 147 (CD147) [reviewed by ^18^].

Recent observations showed that secondary transmission among close household contacts was 53%^19^. Furthermore, recently, the Brazilian media reported that both sibs from 7 pairs of monozygotic adult twins died from SARS-CoV-2 infection within a few days of difference. These observations support the hypothesis that genetic factors, which are largely unknown in SARS-CoV-2, influence infection susceptibility and resistance.

Spouses of infected and symptomatic COVID-19 patients sharing the same bedroom without protective measures represent an efficient approach to identify and ascertain resistant individuals highly exposed to the same viral strain of SARS-CoV-2. Here, we investigated 86 discordant couples, in which one was symptomatic, and the partner remained asymptomatic and serum negative. Since we collected all samples in the first semester of 2020, all couples were likely exposed to the same or closely related viral strains ^20^. Our study suggests that genes of innate and adaptive immune responses may play a vital role in this resistance.

### MHC variants

Analysis of MHC variants showed no important associations between antigen-presentation genes and infection resistance. Unlike our approach, previous studies focusing on HLA genes and SARS-CoV-2 infection, comparing infection outcome (mild versus severe COVID-19) or infection susceptibility in infected individuals against population control samples, show different associated alleles for each population and sample ^4,7,21–24^. Our data indicate that polymorphisms in HLA classical class I and class II have little impact on infection susceptibility. The frequencies of predicted strongest and weakest HLA peptide binders do not differ in discordant couples ^25^. Moreover, in our study, polymorphisms of nonclassical HLA genes involved in immune modulation (e.g., *HLA-E*) were not associated with infection susceptibility in discordant couples. Nevertheless, a higher frequency of HLA-E*01:01 has been reported to be associated with hospitalized patients ^21^, and we cannot rule out the influence of HLA alleles in disease severity.

The only exception of a missense mutation in an antigen presentation gene associated with infection susceptibility was rs9269942/T (mostly within alleles DRB1*04:01 in our sample), which is almost absent in resistant individuals and relatively common in infected ones (Figures 2). The mechanisms underlying this association are not clear. However, HLA-DR molecules are important antigen presenting molecules, essentially for CD4+ T cells, and lower HLA-DRB1 expression has been associated with severity of the SARS-CoV-2 infection^26^. Thus, inappropriate antigen-presentation by some HLA-DR molecules might integrate underlying mechanisms facilitating infection susceptibility. Accordingly, DRB1*04:01 has been reported to be associated with lower HLA-DRB1 mRNA expression than many other alleles^27^. However, this expression profile alone does not explain its association because alleles with lower expression levels are also prevalent in our dataset. DRB1*04:01 is common in European populations (France, Denmark, England, Germany) and relatively rare in Asian and African populations (www.allele-frequencies.net).

Interestingly, most of the hits among the genes that we have studied in the MHC region (Figure 1, upper panel) coincide with two genes, *MICA* and *MICB. MICA* and *MICB* are constitutively expressed in a few cell types, such as fibroblasts and epithelial cells but are markedly upregulated in stress conditions as in cancer and infections^28^. Here, *MICA* variants associated with infection susceptibility are related to higher mRNA expression, while the opposite is observed for *MICB* (supplemental results). MICA and MICB interact with activating receptor C-type lectin-like receptor NKG2D and play an important role in mediating NK and TCD8+ cytotoxic activity ^29^. Previous studies suggest the participation of the unconventional T (uT) cells, gamma-delta T (γδ T), NKT, and NK cells in the immune response against several infections such as tuberculosis, HIV, Influenza A, Influenza A (H1N1) [reviewed by ^30^], as well as SARS-COV-1^31^ and SARS-COV-2 infections^32^. These cells recognize non-peptide antigens in an MHC-independent way and produce mostly inflammatory cytokines such as IFN-γ and eliminate target cells by perforin/granzyme-mediated cytotoxicity in vivo ^33^. Thus, these cells could be a key for a rapid defense against bacterial and viral infections ^34^ and contribute to control viral load through a higher MICB expression (from resistant individuals) activating an effective natural response via NKG2D.

The missense *MICB* variant associated with SARS-CoV-2 infection, MICB*004 and rs3131639/A, is increased in frequency in patients with secondary dengue hemorrhagic fever^35^. Thus, higher MICB expression may positively influence DENV infection control by activating early NKG2D mediated immune responses. A similar mechanism may be important for SARS-CoV-2 infection, and higher MICB expression may play a role in the innate immune defense against SARS-CoV-2. Conversely, variants associated with lower MICB expression, possibly implicating a diminution of NK cytotoxicity, may facilitate infection. Besides, the amino acid exchange (75-N, Table 1) may also be related to ligand/receptor binding impairment or lower protein stability. In our study, we detected that rs3131639/A and MICB*004:01 are related to lower MICB expression (Figure SR3), which is confirmed by the GTEx portal (https://www.gtexportal.org). It is not clear the mechanism underlying this differential expression or whether other polymorphisms in LD with rs3131639 may also play a role in the expression regulation.

*MICA* alleles are also associated with susceptibility to infectious diseases^36^. In the present study, *MICA* variants associated with infection are linked to higher MICA expression (Figure SR4). The mechanism underlying this association is not clear. Other viruses stimulate the release of sMICA to escape NKG2D recognition by activating endocytosis and degradation of the NKG2D receptor ^37^. A recent study suggested that dysregulation of NKG2D-MICA axis could be a possible mechanism of NK cell exhaustion in SARS-CoV-2 infection, resulting in suppressive effects by excessive cytokines in the disease course. SARS-CoV-2 might escape from NKG2D recognition using a similar mechanism of elevated plasma levels of sMICA. For instance, the disintegrin and metalloproteinase17 (ADAM17) activity is upregulated upon binding of SARS-CoV to ACE2, facilitating viral entry. This might be responsible for the higher shedding of MICA after spike-ACE2 interaction during SARS-CoV-2 infection [reviewed by ^38^]. Although not confirmed by protein expression analysis, we can hypothesize that the variants associated with higher MICA expression are also associated with higher sMICA levels and/or NK exhaustion, resulting in NK dysfunction in the early stages of SARS-CoV-2 infection. Indeed, decreased NK cell numbers, impaired cytotoxic activity, but also a biased inflammatory phenotype have been reported in SARS-CoV-2 infection, indicating NK cells likely integrate in the underlying immune dysregulation in COVID-19. In this context, our findings bring additional potential mechanisms involving NK dysfunctions [reviewed by ^39^]. However, only functional assays can confirm this hypothesis.

The susceptibility haplotype formed by the *MICB* variants in Figure 2 is more frequent in Europe (about 23%) and Africa (15%) and less frequent in Asia (9.4%). Conversely, the susceptibility haplotype for MICA (rs2844526/T and rs2596541/C) is more frequent in East Asia (about 72%), South Asia (about 63%), and Europe (about 60%), and less frequent in Africa (about 46%). Interestingly, the *MICB* susceptibility haplotype can be detected in archaic humans, such as the Neanderthal from the Vindija Cave, and the *MICA* susceptibility haplotype in the Neanderthal from the Altai Mountains and one Denisovan.

### LRC variants

We detected no relevant associations between KIR genes and SARS-CoV-2 infection for copy number, SNPs, and predicted protein sequences. Conversely, the vast majority of the hits described in Figures 2 and Table 1 are related to immune modulation and innate immunity, mainly NK cell activation.

*LILRB1* (LIR-1/CD85j/ILT2) and *LILRB2* (LIR-2/CD85d/ILT4) are members of the leukocyte immunoglobulin (Ig)-like receptor (LIR) family within the LRC complex. A previous study indicated that high mRNA expression of LILRB2 might be involved in the progression of SARS-CoV-2 infection^26^. We found missense mutations in the same motifs of LILRB1 (rs31739173/A) and LILRB2 (rs7247025/C) 5-fold more frequently in infected individuals (Figure 2 and Table 1). Our results indicate that LILRB1 and LILRB2 variants associated with higher expression and increased receptor stability are associated with SARs-CoV-2 infection susceptibility. Individuals carrying these variants are prone to higher LILRB1/2 expression, which may downregulate T and NK cell activation by interaction with many receptors, including HLA-G. This possible mechanism shares many similarities with the MICA/MICB pathway, which ultimately down-regulates NK cell cytotoxic activity.

Interestingly, we detected a variant at the leukocyte-associated Ig-like receptor-2 (*LAIR2*, CD306), rs59342471/T, associated with protection against SARS-CoV-2 infection. There is no data for this variant in the GTEx portal, and it is not possible to study its expression in the GEUVADIS dataset because all samples have very low expression of LAIR-2. The human LAIR-1 (hLAIR-1, CD305) is an inhibitory immune receptor expressed on most peripheral blood mononuclear cells and thymocytes, which interacts extensively with collagens, mediating immunoregulatory crosstalk interactions between immune cells and the extracellular matrix microenvironment [reviewed by ^13^]. The inhibitory response of collagens-LAIR-1 interactions may be regulated by LAIR-2, a soluble molecule that competes for collagens and acts as a proinflammatory mediator, resulting in highly activated immune cells^13^.

### Concluding remarks

In short, here we performed a candidate region approach to compare polymorphism in the MHC and LRC regions in COVID-19 infected individuals and exposed partners who were asymptomatic and seronegative for COVID-19. We used a state-of-the-art method to call genotypes and haplotypes in the MHC and LRC. We observed little to no impact of polymorphisms in antigen-presentation genes and KIR genes in resistance or susceptibility to infection. Our results suggest that genes related to immune modulation, mainly involved in NK cell killing activation/inhibition, harbor variants potentially contributing to infection resistance. We hypothesize that individuals prone to produce higher amounts of MICA (possibly soluble), LILRB1, LILRB2, and low amounts of MICB, would be more susceptible to infection. Accordingly, quantitative differences in these NK activity-related molecules could contribute to resistance to COVID-19, likely down regulating NK cell cytotoxic activity in infected individuals but not in resistant partners. Functional assays will provide the means to test the hypothesis of differential NK cell activity between SARS-CoV-2 infected and resistant individuals, involving MICA, MICB and LAIR1/2 molecules.

## Supporting information

Supplementary Results

## Data Availability

Individual level data can be shared by authors upon reasonable request.

https://castelli-lab.net/apps/hla-mapper

https://castelli-lab.net/apps/vcfx

## Supplementary Materials

There are two supplementary files accompanying this manuscript, one for results and other for methods. There are also two tables in EXCEL format, one describing SNP frequencies between groups, and another describing allotype frequencies.

## Data availability

hla-mapper and vcfx are freely available in www.castelli-lab.net/apps/hla-mapper and www.castelli-lab.net/apps/vcfx, respectively. The Phasex program and the Perl script to call HLA alleles from phased VCF are available upon request. GATK is provided by the Broad Institute. SNP and allotype frequencies are available as supplementary files.

## Acknowledgments

The authors are extremely grateful to all volunteers for their participation and collaboration; the nurses for samples collection; the technical team for the material process and data analysis and to the Fleury Laboratory for serology tests. Special thanks to JBS. S.A. for the additional funding.

## Ethics Declaration

This study was approved by the Committee for Ethics in Research of the Institute of Biosciences at the University of São Paulo (CAAE 34786620.2.0000.5464). The informed consent term was obtained from all participants.

## Author information

Conceptualization: ECC, MVC, MSN, DM, MZ

Data curation: MVC, LRBM, MVRS

Formal Analysis: ECC, MSN, MOS, NSBS, HSA, ASS, RNP, CFBC, KN

Funding acquisition: MZ

Investigation: MVC, MSN, MOS, ECN, KSS

Methodology: ECC, MSN, MOS, CTMJ, DM, KN

Project administration: MZ

Software: ECC, MSN, MOS

Visualization: ECC, MSN, MOS, CTMJ, KN

Writing – original draft: ECC, MVC, MSN, MOS

Writing – review & editing: NSBS, HSA, ASS, RNP, CFBC, CTMJ, DM, KN, LRBM, MVRS, JE, VRC, RHB, MHH, JYM, ECN, VC, KSS, MLCM, JK, MMN, RMBM, MRPB, MZ

