## Supplementary Methods for "“Immunogenetics of resistance to SARS-CoV-2 infection in discordant couples”"

**Summary**

**Volunteers Recruitment and datasets**	**2**

**Exome sequencing, variant call, and variant refiniment**	**3**

**Ancestry assessment**	**3**

**MHC and KIR mapping optimization, genotyping, and haplotyping**	**3**

Figure SM1: The workflow used in this study, from the raw sequencing data to complete sequences for HLA and KIR genes. We used the datasets marked in red for the SARs-CoV-2 association analyses.	5

**Getting the name of HLA and KIR alleles**	**5**

**Evaluation of copy number variation in the LRC**	**6**

Figure SM2: Histogram of the ratios between read depth at every exon at KIR2DL3 and the read depth at every exon on the TNF and LTB loci, with the cutoffs for the absence of KIR2DL3, the presence of a single copy, or the presence of two copies.	6

**Statistical analyses for the association study**	**6**

**Expression analysis in the GEUVADIS dataset**	**7**

**Evaluation of the susceptibility markers in archaic humans**	**8**

**References**	**8**

### Volunteers Recruitment and datasets

The couple's infected members were divided into subgroups based on their COVID-19 clinical condition, using the severity scales proposed by World Health Organization (WHO-2019-nCoV-clinical-2020.5), and elaborated by Gandhi and collaborators ^1^ . The subgroups are: (a) Asymptomatic, with no presence of symptoms; (b) mild illness, with the presence of most common symptoms such as fever, cough, dysfunction of smell (anosmia) and taste (dysgeusia) but no shortness of breath (dyspnea), and no hospitalization requirement; (c) moderate illness, with the presence of most common symptoms including dyspnea and clinical or radiographic evidence of lower respiratory tract disease but no hypoxemia (blood oxygen saturation of 94% or higher), in which hospitalization may be required; and (d) severe illness, with the presence of most common symptoms, dyspnea, hypoxemia, pulmonary impairment, and hospitalization in an intensive care unit.

The infected member had COVID-19 confirmed by RT-PCR, and they were asymptomatic (3.2%) or had mild (78.7%), moderate (11.7%), or severe (6.4%) symptoms.

For sample collection, 6 mL of whole blood (WB) from each partner was collected after venipuncture using the BD Vacutainer tube system with ethylenediaminetripotassium (BD Catalog. 360057) for DNA extraction. DNA was automated extracted using the QIAsymphony® robot (Qiagen) with elution volumes of 200μL and following the protocols as recommended by the manufacturer. DNA quality was assessed with NanoDrop® (Thermo Fisher Scientific) and concentration with Qubit® (Thermo Fisher Scientific).

We also compared the resistant and the infected groups with a population-based sample from the same city ^2^. For this purpose, we resampled the SABE cohort using an in-house Perl script selecting 5 individuals of the same sex and similar ancestry background for each resistant (SABE subsample, group C) and infected (SABE subsample, group D) individuals. Thus, both these SABE subsamples are paired with the resistant or infected groups by ancestry and sex. The overlap between both SABE subsamples is 54.6%.

Table SM1: The description of the datasets and groups used to evaluate SARS-CoV-2 infection susceptibility and resistance.

| **Group** | **Description** | **Size** | **Mean age** | **Sex (%Male)** | **European Ancestry** | **African Ancestry** | **Asian Ancestry** | **Native American Ancestry** |
| --- | --- | --- | --- | --- | --- | --- | --- | --- |
| Group A | Resistant | 86 | 45.8 | 33.72 | 85.96% | 6.17% | 1.83% | 5.95% |
| Group B | Infected | 86 | 46.8 | 61.62 | 83.05% | 8.23% | 1.05% | 7.67% |
| Group C | SABE subsample paired to Resistant | 430 | 71.0 | 33.72 | 85.67% | 6.38% | 2.32% | 5.58% |
| Group D | SABE subsample paired to Infected | 430 | 71.0 | 61.6 | 83.23% | 8.72% | 1.19% | 6.79% |

### Exome sequencing, variant call, and variant refiniment

Reads were aligned to human reference GRCh38 using the run-bwamem algorithm from bwa.kit 0.7.15 package (https://github.com/lh3/bwa/tree/master/bwakit). After alignment, we used Picard tools 2.18.7 (http://broadinstitute.github.io/picard) to mark duplicates and GATK 4.0.9 ^3^ to perform Base Quality Score Recalibration (BQSR, BaseRecalibrator tool). Following GATK's Best Practices for germline short variant discovery ^4^ and using the GATK 4.0.9 tools, we generated individual GVCFs using HaplotypeCaller (GVCF mode). We then combined the GVCFs of all individuals with CombineGVCFs to jointly call variants using GenotypeGVCFs and perform Variant Quality Score Recalibration (VQSR, VariantRecalibrator). The mean depth of coverage for the 172 individuals was 95X, ranging from 75 to 150X.

We checked the biological sex of individuals against informed sex using the coverage ratio between the X and Y chromosomes. We used PC-Relate implemented in the GENESIS R package to check for relatives or contamination in the sample ^5^. No individual failed sex or kinship checks.

### Ancestry assessment

We used our previously developed CEGH-Filter to evaluate the quality of called variants and genotypes ^2^. For analysis, we removed GATK non-PASS, CEGH, non-vSR, and multiallelic variants.

Ancestry was inferred in ADMIXTURE v1.3 ^6^, based on likelihood models and the information about allele frequencies of the different parental populations. As parental populations, we used samples from the 1000 Genomes project ^7^ and HGDP-CEPH ^8^ totalizing 602 African, 624 European, 630 East Asian, and 118 Native American, with more than 95% of ancestry inferred for that ethnic group, according to the analysis unsupervised (K = 4) in ADMIXTURE. Ancestry inferences were performed with supervised analysis (K = 4) after applying linkage disequilibrium filters (*r^2^* = 0.1) within a sliding window of 50Kb and a shift step of 10Kb, totaling 50,995 SNPs. Lastly, we retrieved the proportion of each ancestry (European, African, Native American, and East Asian) for each sample, which was used as covariates in subsequent analyses.

### MHC and KIR mapping optimization, genotyping, and haplotyping

The workflow used in this study is illustrated in Figure SM1. First, we aligned reads to the human reference GRCh38 using the run-bwamem algorithm from bwa.kit 0.7.15 package (<https://github.com/lh3/bwa/tree/master/bwakit>). The SAM file produced by using BWA was converted to a BAM file, sorted, and indexed using Samtools ^9^. This BAM file is the input for the HLA and KIR alignment optimization by using hla-mapper.

We used hla-mapper version 4.0.12 ([www.castelli-lab.net/apps/hla-mapper](http://www.castelli-lab.net/apps/hla-mapper)) to optimize alignments in the MHC and KIR clusters ^10^, using a similar approach as previous manuscripts addressing HLA and KIR genes in Brazil ^2,11–13^. Each of these clusters are prone to alignment and genotyping errors ^10,14^, and there is a different hla-mapper database used for each cluster that must be downloaded in the developers’ website. This procedure minimizes the alignment errors, allowing a more accurate genotyping and haplotyping procedure. The end product of such optimization is a BAM file for each sample.

For variant calling, we used GATK HaplotypeCaller in the GVCF mode ^4^. After converting the GVCF files to VCF by using GATK GenotypeGVCFs, we recoded the VCF file using vcftools to correct minor encoding errors ^15^. After, we proceeded with a variant refinement step by first using vcfx ([www.castelli-lab.net/apps/vcfx](http://www.castelli-lab.net/apps/vcfx)) algorithm checkpl (genotype likelihood = 0.95), which introduces missing alleles in genotypes with low likelihood (for example, an homozygous genotype in a region with only two reads, or an heterozygous genotype in which 90% of all reads point to the same nucleotide), and second with algorithm evidence (default parameters), which annotates each variant with quality control parameters (number of heterozygous sites with even distribution of reads per allele, number of homozygous genotypes for alternative alleles, and others). Then, we used vcfx filter to recover only variants annotated as PASS or WARN in the previous steps. The final step for variant refinent is the removal of alternative alleles that are no longer present in the dataset, recoding the VCF file by using *Bamtools view* --trim-alt-alleles --min-ac 1 ^16^. After, we filtered variants that coincided with the regions that are captured by the IDT xgen-V1, with a tolerance of 100 nucleotides upstream and downstream each region. We also filtered the variants that coincide with the regions optimized by hla-mapper (Figure 1). The end-product of these procedures is an unphased VCF file (Figure SM1) that is used for the association analysis described later, and also used as input for the haplotyping procedure.

To call haplotypes, we combined read-aware phasing and probabilistic models. First, we converted multi-allelic variants into bi-allelic ones using *Bamtools norm*. Then, we used GATK ReadBackedPhasing (from GATK 3.8) to infer the physical phasing between closely related variants. Similar results can be obtained with WhatsHap ^17^. This step can be parallelized using a Perl script to speed up the process, since neither the ReadBackedPhasing nor WhatsHap are multithreaded. This script is available upon request. The VCF file containing the phased data is the input for the haplotyping procedure using a local program named phasex.

Phasex is a program that automates multiple haplotyping runs by using Shapeit 4 ^18^ and the data obtained by ReadBackedPhasing or WhatsHap. It performs many independent runs, comparing the results afterwards, fixing haplotypes for a sample in which at least 95% of the runs indicate the same results. Then, this step is repeated many times until the number of samples with the same haplotype at least 95% of the runs do not increase any further. The end product of this procedure is phased bi-allelic VCF, which is then converted to multi-allelic variants using *bamtools norm*. It should be mentioned that we have evaluated each MHC/KIR gene separately by retrieving the variants corresponding to each locus, and running phasex for each gene. For KIR genes, we first calculated the copy number of each locus (detailed later), and then we removed all samples that do not carry the target gene before the haplotyping procedure. Phasex is available upon request. The missing alleles introduced by vcfx are imputed by Shapeit 4.

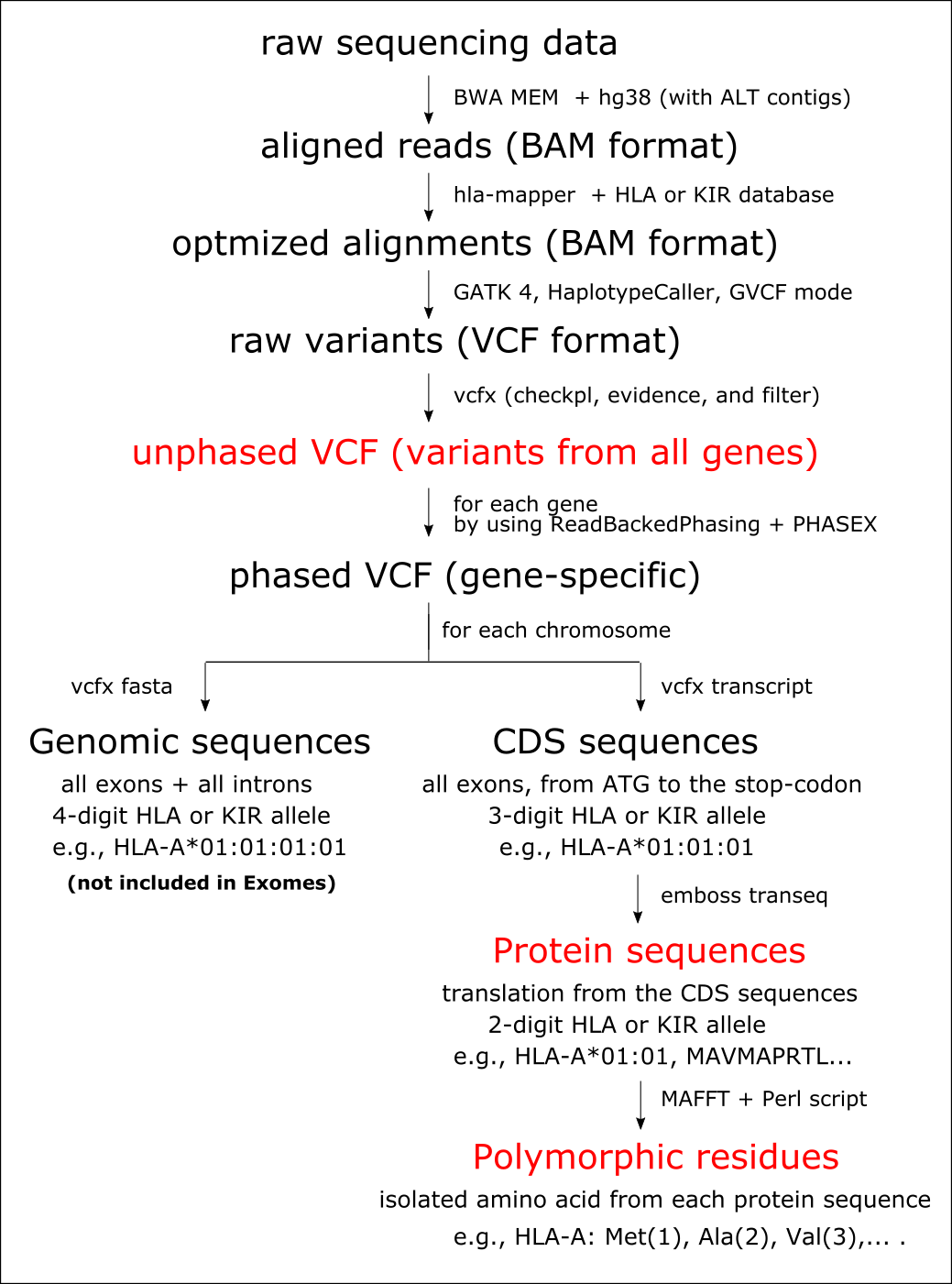

#### Figure SM1: The workflow used in this study, from the raw sequencing data to complete sequences for HLA and KIR genes. We used the datasets marked in red for the SARs-CoV-2 association analyses.

### Getting the name of HLA and KIR alleles

Using a Perl script to automate the process, we first exported the phased VCF to complete sequences (only exons) of each gene. To do that, we used vcfx transcript, indicating chromosome 6 or 19 as a reference, the phased VCF, and a BED file with coordinates of each exon (starting from the first translated ATG). This procedure produces two sequences per individual, one for each chromosome. Then, we detected each different sequence, comparing them with the ones available in the IPD-IMGT/HLA and IPD-IMGT/KIR database ^19^. When the sequence we detected was found in the database, we updated its name accordingly. When it was a new sequence, we named the sequence as a new one (Figure SM1).

After, we translated the sequence of each chromosome using emboss transeq, generating a fasta file with the proteins encoded by each chromosome of each individual. Then, we detected each different sequence, also naming them following the ones in the IPD-IMGT database, or identifying them as new sequences. All these steps are automated by the Perl script (available upon request), which generates fasta files as described above, and a final database indicating the alleles of each individual considering the exonic sequence and the protein sequence (Figure SM1). All samples presenting just one copy of a specific KIR gene presented two copies of the same exonic or protein sequence, and one of these copies were manually substituted to "null allele".

### Evaluation of copy number variation in the LRC

To calculate the copy number of each KIR gene, first we calculated read depth in all exons of a KIR gene, and also in all exons from genes *TNF* and *LTB* on chromosome 6, by using *samtools coverage* and indicating the coordinates of each exon. We calculated the average read depth in TNF/LTB (the reference) and in each KIR gene (the query), and the ratio query/reference. This procedure was automated by using a Perl script. After, we plotted the ratios in a histogram with bin width of 0.02, observing the distribution to manually set the thresholds for an absence of a gene, one copy, two copies, three copies, etc, assuming that all individuals present two copies of the reference. Figure SM2 demonstrates this calculation for gene *KIR2DL3*, with the cutoffs established manually by observing the histogram distribution.

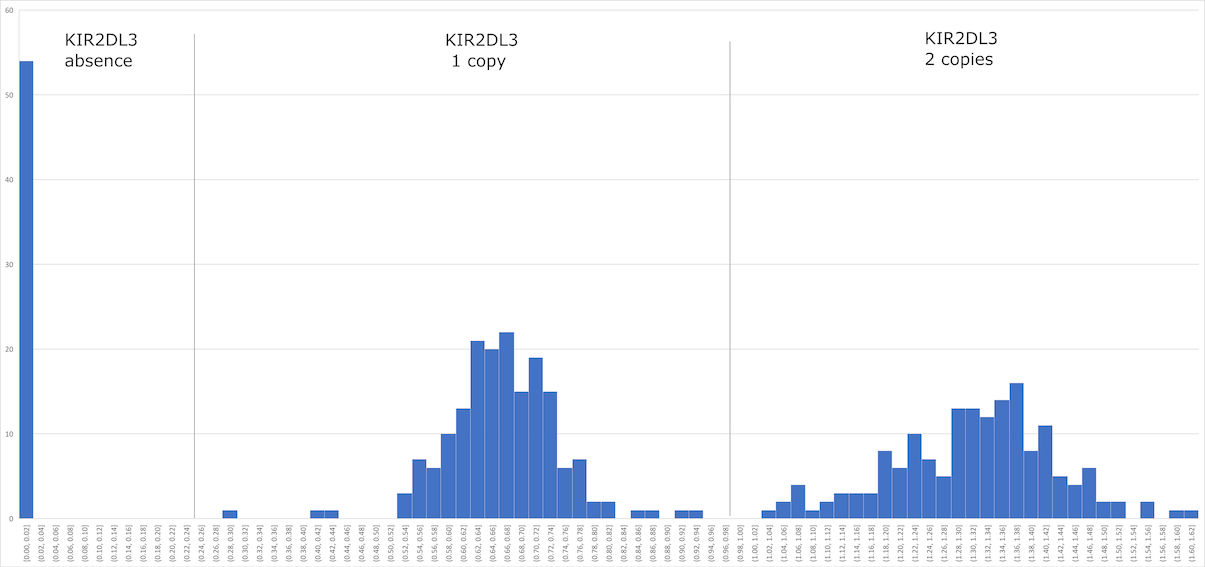

#### Figure SM2: Histogram of the ratios between read depth at every exon at *KIR2DL3* and the read depth at every exon on the *TNF* and *LTB* loci, with the cutoffs for the absence of *KIR2DL3*, the presence of a single copy, or the presence of two copies.

### Statistical analyses for the association study

First, we tested for the association between bi-allelic and multi-allelic variants and two phenotypes: susceptibility or resistance to infection. Because there are many multi-allelic variants, to perform the logistic regression, we used a local Perl script to convert the unphased VCF file (Figure SM1) to a plink-like table in which each allele of a variant is considered an independent marker. Genotypes carrying missing alleles are disregarded by registering "NA" to the presence of this marker. Then, we used R to fit a regression model for each marker, controlling for sex, age, and ancestry. This approach is not easily applicable for the KIR genes because a marker might be absent due to a different nucleotide sequence or due to gene deletions. Because of that, this approach was applied only for the genes in the MHC complex that are optimized by hla-mapper (Figure 1A) and the ones in the LRC not presenting copy number variation (*KIR2DL4, KIR3DL2, KIR3DL3, LAIR1, LAIR2, LILRB1*, and *LILRB2*). We filtered all markers that presented a P-value lower than 1%, summarizing these markers and their frequencies in Figure 2. We also plotted the variants for the MHC in a Manhattan plot, by using R and library qqman, and highlighting variants with P-values lower than 1% (Figure SR1).

After translating the exonic sequences to proteins (Figure SM1), we explored whether specific protein sequences within genes from the MHC and the LCR are associated with the phenotypes. We used a similar Perl script to create a plink-like table that considers each different allotype as an independent marker, and indicates the number of copies of these markers for each sample. We tested the frequency of each different protein sequence (the allotypes) among groups by using logistic regression and R, also controlling for sex, age, ancestry. We filtered all protein sequences that presented a P-value lower than 1% in each test, summarizing them in Table 1. We also applied a correction for multiple tests within each gene, by multiplying the *P*-value for a protein sequence by the number of different proteins observed for that particular gene.

Because more than one allele in multi-allelic variants may encode the same amino acid, and because multiple full-length proteins may present the same amino acid in one specific position, we also tested the frequency of every amino acid. First, for each locus, we aligned the complete protein sequences of every individual (two per individual) by using MAFFT (Figure SM1). After, we used a Perl script that created a plink-like table with every allele (different amino acid) in every position as an independent marker, and the number of copies of this marker in each sample. We fitted a logistic regression by using R, also controlling for sex, age, ancestry. We filtered all amino acids/positions that presented a *P*-value lower than 1%, summarizing them in Table 1. We also applied a correction for multiple tests within each gene, by multiplying the *P*-value for an amino acid/position by the number of polymorphic positions within the locus.

### Expression analysis in the GEUVADIS dataset

For some variants associated with SARs-CoV-2 infection susceptibility and COVID-19 severity, we investigated whether these variants influence the expression of their respective genes. We used the GEUVADIS dataset of RNA-seq data from 460 samples from the 1000 Genomes project ^20^ to evaluate the expression levels. First, we genotyped all samples in the new 1000 Genomes release of high-coverage sequencing ^21^, by applying hla-mapper to optimize the BAM files available for download and applying the same pipeline to call and refine genotypes (Figure SM1).

Second, we aligned the raw sequencing data from GEUVADIS using STAR ^22^ and the human genome draft as reference (version hg38 without ALT contigs), considering all known transcripts annotated for the hg38 genome. Then, we applied a beta version of the hla-mapper suitable for RNA-seq data (on development), that optimizes the alignment of the same genes as for the DNA version. With these optimized alignments, we used subread featureCounts ^23^ to count the number of reads aligned in each gene. We used the total number of aligned reads and the length of each gene as indicated by subread to calculate TPM for each locus.

Third, for each tested variant, by using a local Perl script, we tracked the TPM values for individuals carrying different genotypes, producing a text file composed of the sample name, its genotype, and the TPM value for a specific locus. By using R, we compared the distribution of TPM values among individuals carrying different genotypes (using wilcoxon or t-test), plotting this distribution in boxplots.

### Evaluation of the susceptibility markers in archaic humans

We have downloaded the aligned BAM files from two Neanderthals from the Altai Mountains ^24^ and the Vindija Cave in Croatia ^25^, and one Denisovan ^26^. Then, we applied hla-mapper to optimize the alignment in the MHC region. By using Integrative Genome Viewer (https://software.broadinstitute.org/software/igv/), we manually evaluated each associated position described in Figure 2 (upper panel).
