## Supplementary Results for "“Immunogenetics of resistance to SARS-CoV-2 infection in discordant couples”"

**Summary**

**Figure SR1
Manhattan plot for association between variants across the Major Histocompatibility Complex.**	**2**

**Figure SR2
Frequency of associated variants in the MHC and LRC in infected and resistant individuals, and in the general population.**	**3**

**Potential mechanisms underlying the associations**	**4**

MICB*004 and rs3131639	4

Figure SR3: MICB mRNA expression profile for individuals from the GEUVADIS cohort presenting different genotypes for rs3131639.	4

MICA, variants rs2844526, rs2596541, and rs2252223	4

Figure SR4: MICA mRNA expression profile for individuals from the GEUVADIS cohort presenting different genotypes for rs2844526.	5

DOB*01:02 and rs2071554	5

LILRB1 and LILRB2, missense variants	6

**Table SR1
Proportion of individuals in the SARS-CoV-2 Resistant and Infected groups carrying at least a copy of each KIR gene**	**6**

**Table SR2
SNP frequencies in SARS-CoV-2 Resistant and Infected individuals**	**6**

**Table SR3
Allotype frequencies in SARS-CoV-2 Resistant and Infected individuals**	**7**

**References**	**7**

### Figure SR1 Manhattan plot for association between variants across the Major Histocompatibility Complex.

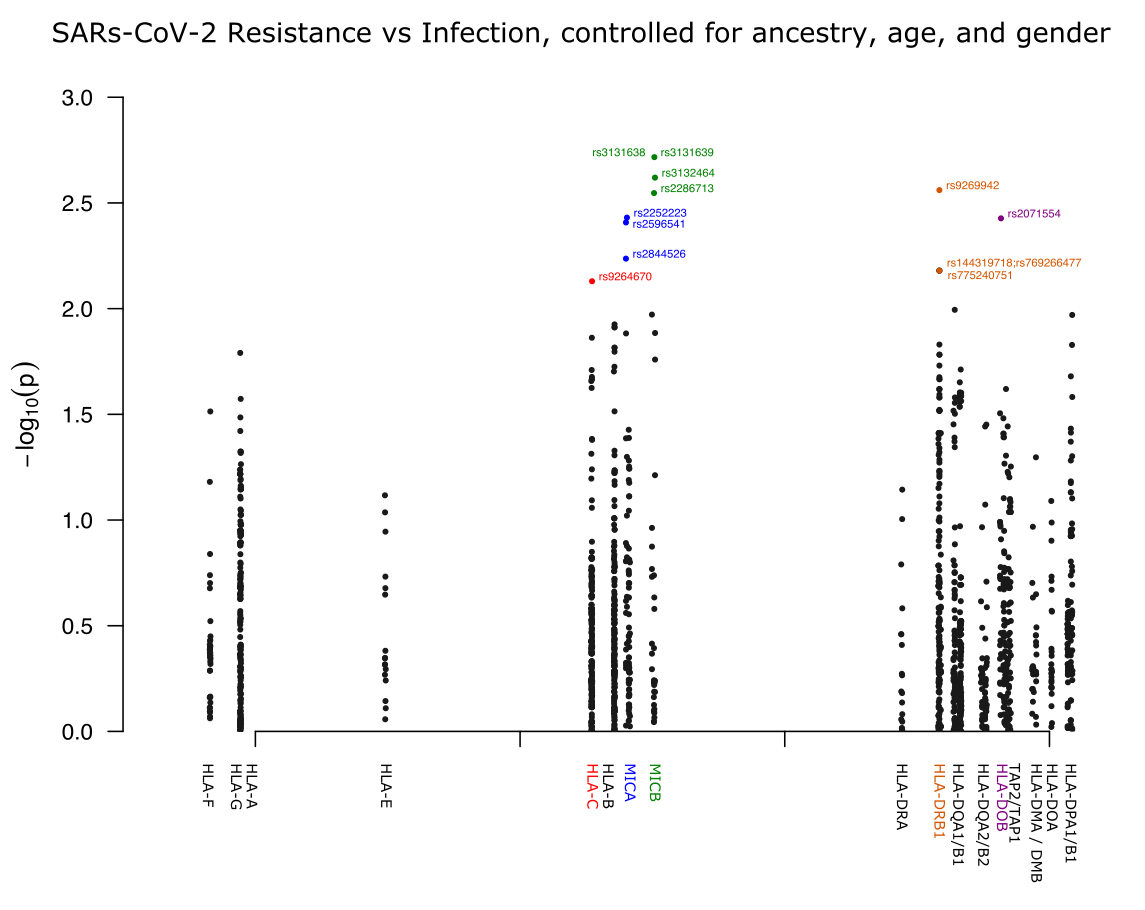

Figure SR1: Manhattan plot for association between bi-allelic and multi-allelic variants across the Major Histocompatibility Complex region on chromosome 6, and the phenotypes susceptibility or resistance to SARs-CoV-2 infection, controlled for ancestry, sex, and age. Variants with *P*-values < 0.01 are highlighted.

### Figure SR2 Frequency of associated variants in the MHC and LRC in infected and resistant individuals, and in the general population.

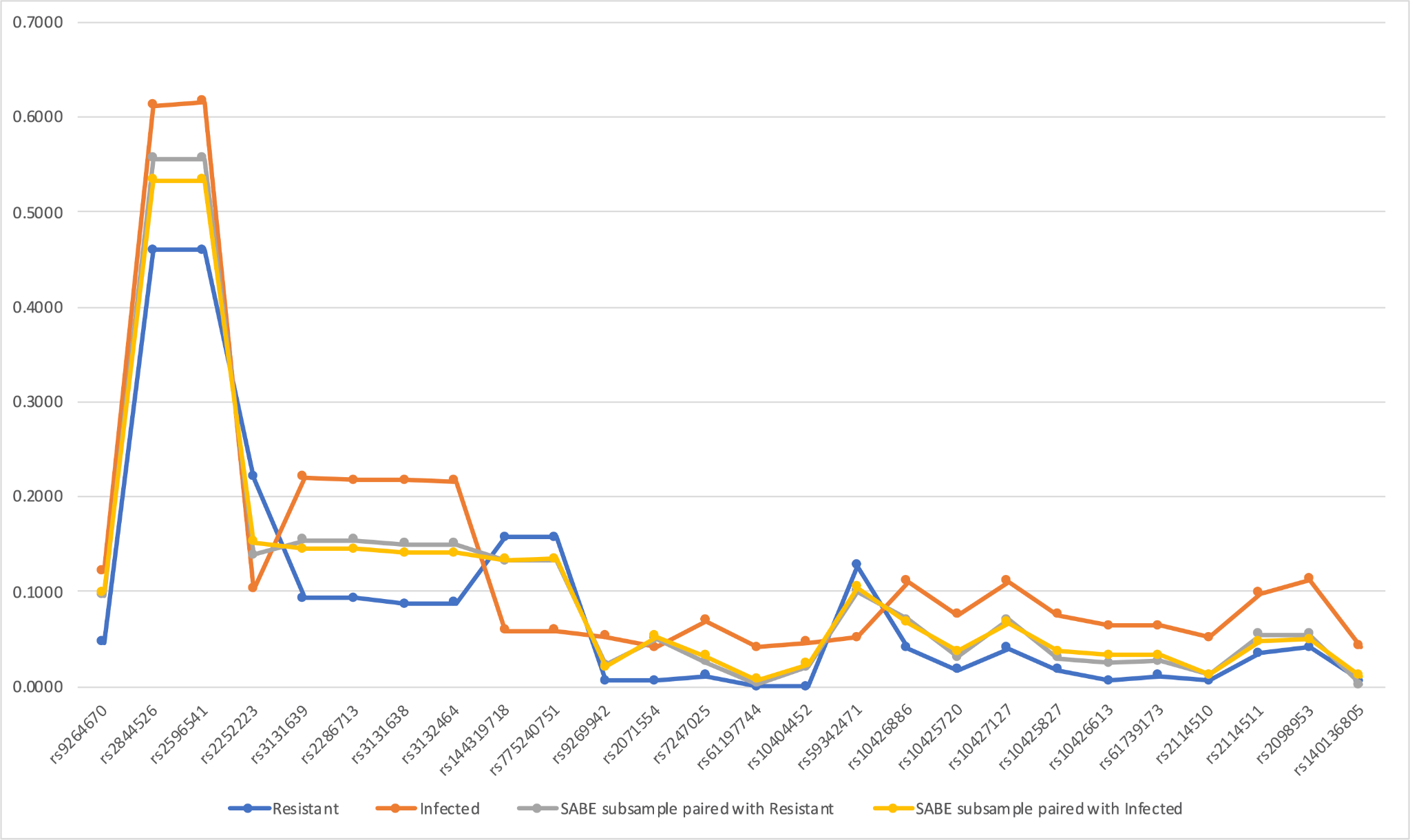

Figure SR2: The frequency of alleles listed in Figure 2 for the MHC and LRC regions, in four different groups: the SARS-CoV-2 resistant individuals (blue), SARS-CoV-2 infected individuals (orange), in a population-based sample paired with resistant individuals by genetic ancestry and sex (gray), and in a population-based sample paired with infected individuals by genetic ancestry and sex (yellow).

### Potential mechanisms underlying the associations

#### MICB*004 and rs3131639

*MICB* encodes a stress-induced molecule (e.g., during infection diseases) that is a ligand for the NKG2D type II receptor, modulating cytotoxicity of T and NK cells. rs3131639 reference allele (Adenine) encodes Asn (neutral, polar) while the alternative allele Guanine encodes Asp (acid, charged). rs3131639/A defines the allele MICB*004:01. Allele rs3131639/A is associated with lower *MICB* mRNA expression according to the GTEx portal. We evaluated the expression profile of 438 individuals from the GEUVADIS dataset ^1^, and rs3131639/A was also associated with lower levels of mRNA (Figure SR3). It is not clear the mechanism underlying these expression levels since none of these SNPs seem to coincide with regulatory elements ^2^. Thus, we can hypothesize that individuals carrying MICB*004/rs3131639A are prone to express less MICB than non-carriers, thus activating less NK cells via NKG2D, facilitating infection.

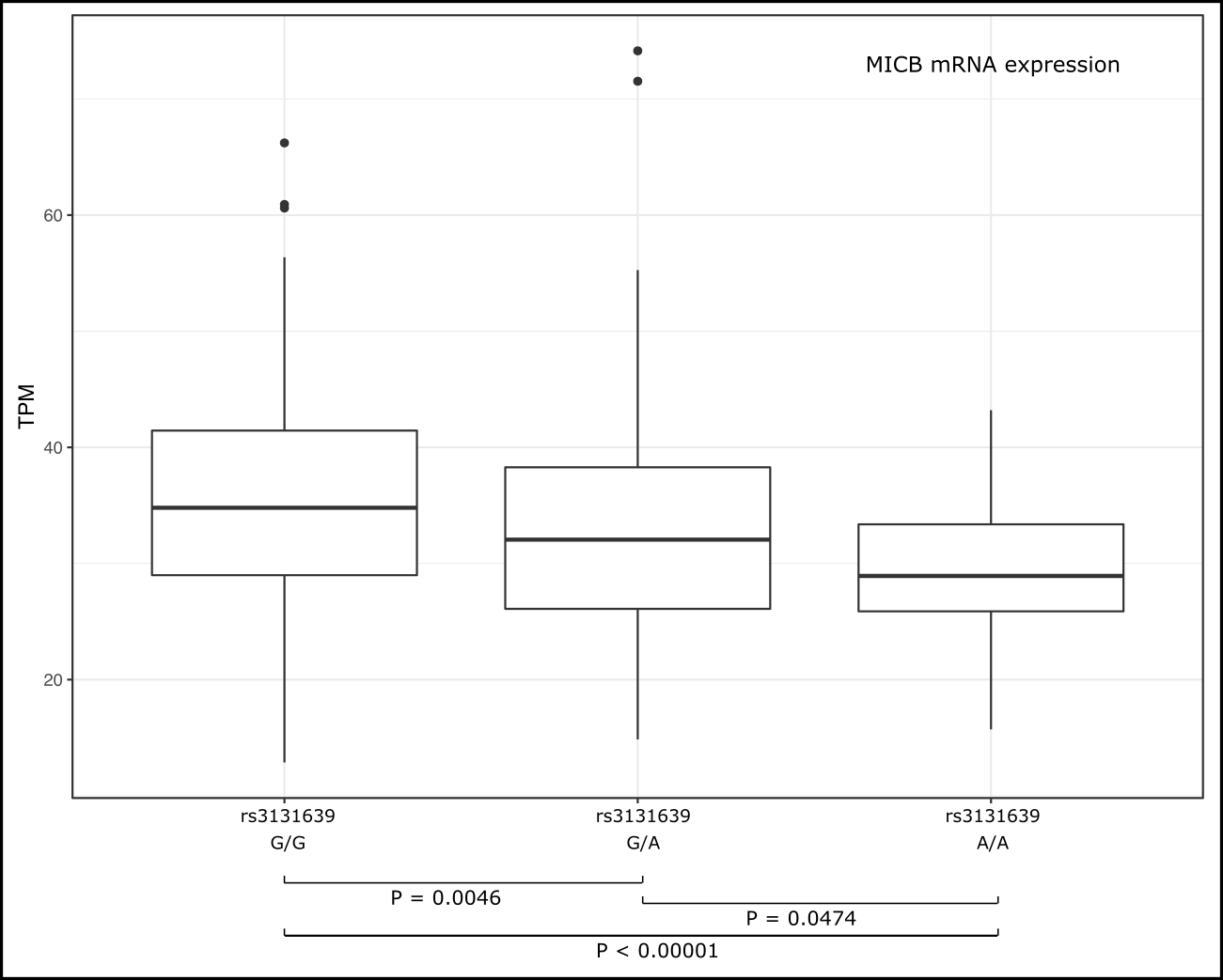

##### Figure SR3: MICB mRNA expression profile for individuals from the GEUVADIS cohort presenting different genotypes for rs3131639.

#### *MICA, variants* rs2844526, rs2596541, and rs2252223

*MICA* is also induced by stress and is a ligand for the NKG2D. We detected two *MICA* variants associated with infection susceptibility, rs2844526 and rs2595541, both located more than 3.5 Kb upstream the *MICA* first translated ATG. These variants are eQTLs for *MICA* expression in the GTEx portal, with the susceptibility alleles (Figure 2) associated with higher MICA mRNA expression levels in many tissues, including lung. Both these variants lay on a Candidate cis-Regulatory Element (cCRE), EH38E3459116, with a proximal enhancer-like signiture, and a CpG island of 440 nucleotides ^2^. The association between rs2844526/T and rs2596541/C and higher *MICA* mRNA expression levels was confirmed in the GEUVADIS dataset, Figure SR4.

We also detected a protective allele, rs2252223/C, but this variant does not influence MICA expression according to GTEx and in the GEUVADIS dataset. However, there is also a candidate Cis-regulatory element in this region, EH38E2459125, and a CpG island ^2^.

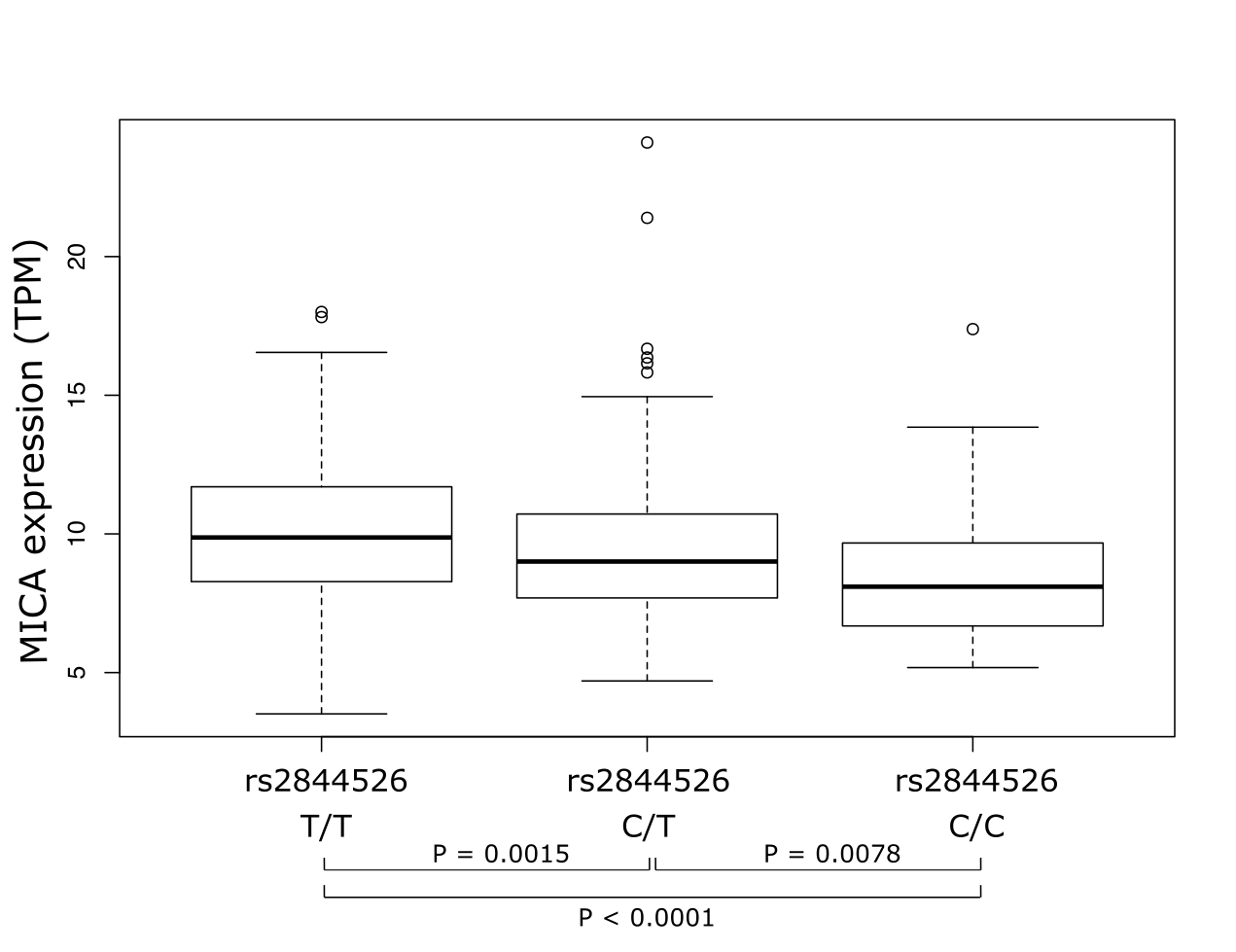

##### Figure SR4: MICA mRNA expression profile for individuals from the GEUVADIS cohort presenting different genotypes for rs2844526.

#### DOB*01:02 and rs2071554

HLA-DO is an heterodimer formed by HLA-DOA and HLA-DOB, and it is usually located in intracellular vesicles anchored to the membrane. Variant rs2071554/T is a missense variation that results in an arginine to glutamine substitution in the HLA-DOB signal peptide, at position 18 (R18Q). Polyphen2 analysis ^3^ predicted this amino acid exchange as possible damaging to the protein function (Polyphen2: 0.897, sensitivity: 0.82, specificity: 0.94). Similarly, SIFT ^4^ predicted this exchange to be deleterious (SIFT score: 0.016). However, the impact of this exchange in protein function is not clear. Besides the modification in the signal peptide, it is unclear if this variant influences HLA-DOB cellular localization and expression. While SignalP-5 ^5^ predicts that both signal peptides (the most frequent one and the one carrying 18Q due to rs2071554) as secretory signal peptides, PrediSi (<http://www.predisi.de>) predicts only the mutated one as a secretion peptide. Therefore, it is possible that cellular localization is influenced by this mutation. According to GTEx and our analysis of the GEUVADIS dataset (*P* = 0.0643), this variant does not influence HLA-DOB expression levels. However, the region encompassing rs2071554 presents a cCRE with a promoter-like signature ^2^.

#### LILRB1 and LILRB2, missense variants

Variants rs61739173/A (LILRB1, G350R) and rs7247025/C (LILRB2, R349G) occur in the same D3/D4 domain motif. LILRB1 exchange G350R is associated with less flexibility and higher protein stability, while LILRB2 R349G seems to not affect protein stability ^6^. There are no regulatory elements that coincide with any of the associated LILRB1/LILRB2 variants ^2^. However, the GTEx portal indicates that rs10426886/A, rs10427127/C, and rs2098953/C, and variant rs2114511/C, are all associated with higher expression of LILRB1 in the lung, and much higher LILRB2 expression in EBV-transformed lymphocytes. Likewise, heterozygosis for the *LILRB2* variants rs7247025/C and rs10404452/A are associated with higher LILRB2 expression in many tissues. All these variants are overrepresented among infected individuals.

### Table SR1 Proportion of individuals in the SARS-CoV-2 Resistant and Infected groups carrying at least a copy of each KIR gene

| **Gene** | **Resistant (N = 86)** | **Infected (N = 86)** | ***P*-value *** |
| --- | --- | --- | --- |
| KIR2DL1 | 0.9535 | 0.9535 | 0.7099 |
| KIR2DL2 | 0.5814 | 0.5930 | 0.8648 |
| KIR2DL3 | 0.8605 | 0.8721 | 0.9872 |
| KIR2DS1 | 0.4767 | 0.5000 | 0.5236 |
| KIR2DS2 | 0.6047 | 0.6047 | 0.9116 |
| KIR2DS3 | 0.3721 | 0.3488 | 0.7972 |
| KIR2DS4 | 0.9419 | 0.8953 | 0.3287 |
| KIR2DS5 | 0.3140 | 0.4186 | 0.0845 |
| KIR3DL1 | 0.9535 | 0.8953 | 0.2118 |
| KIR3DS1 | 0.4419 | 0.4767 | 0.5301 |

* controlled for sex, age, and genetic ancestry

### Table SR2 SNP frequencies in SARS-CoV-2 Resistant and Infected individuals

Table SR2 lists the frequencies of each variant in the MHC and LRC region between infected individuals and resistant ones, and the strength of association (P-value) when controlling for ancestry, sex, and age. This table is available as a spreadsheet in EXCEL format (SNP_frequency.xlsx).

### Table SR3 Allotype frequencies in SARS-CoV-2 Resistant and Infected individuals

Table SR3 lists the allotypes for each gene, their frequencies, and the strength of association (P-value) when controlling for ancestry, sex, and age. The absence of a gene copy (due to gene deletion - copy number variation) is registered as allele "null". For allotypes that resemble one already reported with minor modifications, the allotype name is followed by "_comp". This table is available as a spreadsheet in EXCEL format (Allotype_frequency.xlsx).
